# Quantitative prediction of right ventricular and size and function from the electrocardiogram

**DOI:** 10.1101/2023.04.25.23289130

**Authors:** Son Q. Duong, Akhil Vaid, Ha My Thi Vy, Liam R. Butler, Joshua Lampert, Robert H. Pass, Alexander W. Charney, Jagat Narula, Rohan Khera, Hayit Greenspan, Bruce D. Gelb, Ron Do, Girish Nadkarni

## Abstract

**Background:** Right ventricular ejection fraction (RVEF) and end-diastolic volume (RVEDV) are not readily assessed through traditional modalities. Deep-learning enabled 12-lead electrocardiogram analysis (DL-ECG) for estimation of RV size or function is unexplored.

**Methods:** We trained a DL-ECG model to predict RV dilation (RVEDV>120 mL/m^2^), RV dysfunction (RVEF≤40%), and numerical RVEDV/RVEF from 12-lead ECG paired with reference-standard cardiac MRI (cMRI) volumetric measurements in UK biobank (UKBB; n=42,938). We fine-tuned in a multi-center health system (MSH_original_; n=3,019) with prospective validation over 4 months (MSH_validation_; n=115). We evaluated performance using area under the receiver operating curve (AUROC) for categorical and mean absolute error (MAE) for continuous measures overall and in key subgroups. We assessed association of RVEF prediction with transplant-free survival with Cox proportional hazards models.

**Results:** Prevalence of RV dysfunction for UKBB/MSH_original_/MSH_validation_ cohorts was 1.0%/18.0%/15.7%, respectively. RV dysfunction model AUROC for UKBB/MSH_original_/MSH_validation_ cohorts was 0.86/0.81/0.77, respectively. Prevalence of RV dilation for UKBB/MSH_original_/MSH_validation_ cohorts was 1.6%/10.6%/4.3%. RV dilation model AUROC for UKBB/MSH_original_/MSH_validation_ cohorts 0.91/0.81/0.92, respectively. MSH_original_ MAE was RVEF=7.8% and RVEDV=17.6 ml/m^2^. Performance was similar in key subgroups including with and without left ventricular dysfunction. Over median follow-up of 2.3 years, predicted RVEF was independently associated with composite outcome (HR 1.37 for each 10% decrease, p=0.046).

**Conclusions:** DL-ECG analysis can accurately identify significant RV dysfunction and dilation both overall and in key subgroups. Predicted RVEF is independently associated with clinical outcome.

## Introduction

Right ventricular size and functional metrics have important prognostic implications for many diseases including cardiomyopathy, pulmonary hypertension, and structural heart disease^1^. Volumetric measurements such as RV ejection fraction (RVEF) and end diastolic volume (RVEDV) are correlated with major adverse outcomes^2^ and have particular importance in patients with congenital heart disease^3^ and pulmonary hypertension^4^.

However, current tools for accurate quantification of RVEF and RVEDV are limited. Estimation of volumes by traditional two-dimensional echocardiographic RV measurements are not recommended in adults or children^5,6^ due to poor reproducibility, especially in the setting of significant pathology^7^. Three-dimensional echocardiography is a promising novel technology, but has significant technical and processing limitations and is not broadly available^8,9^. Cardiac MRI (cMRI) is the clinical reference standard for volumetric quantification, but cMRI is not globally accessible, is time- and resource-intensive, and cannot be performed in some patients such as those with implanted devices and those who will not tolerate the exam.

There is an urgent clinical need for novel methods to assess RV size and function which are validated against gold-standard metrics, widely available, and simple to perform. We sought to develop such a method through application of modern machine learning techniques to a ubiquitous clinical tool: the 12-lead electrocardiogram (ECG).

Recent developments in deep learning (DL) technology have produced models to interpret ECG for accurate diagnosis of structural heart disease and prognosis^10^, but the use of DL on ECG to quantify RVEDV and RVEF has not been explored. We hypothesized that we could develop a DL-algorithm to accurately quantify RVEDV and RVEF from a pathological clinical cohort and a large healthy registry of paired ECG and cMRI.

## Methods

### Model Development

Broadly, we trained a convolutional neural network on a larger dataset of paired ECG and cMRI data from the UK Biobank (UKBB), and then fine-tuned the model on a smaller retrospectively collected cohort of clinically indicated cMRI-ECG pairs from a single urban referral center. We then performed a prospective validation on the subsequent four months following the model training/testing set. This study was reviewed and approved by the Institutional Review Board of the Icahn School of Medicine at Mount Sinai.

### UK Biobank dataset

We accessed paired ECG and cMRI data from the UK Biobank. Participant enrollment and cMRI acquisition parameters are described previously^11^. We utilized a previously validated automated segmentation method to obtain RVEDV and RVEF^12^ from the b-SSFP cine short axis stack. This method contours the right ventricle to include the papillary muscles within the RV volume. We excluded entries with RVEF values < 10% or > 80%. Per the UKBB study protocol, a 12-lead ECG was obtained at the same visit as cMRI. ECG acquisition parameters are previously published^13^, and were accessed in XML format with preprocessing per below.

### Clinical Dataset

As the UK Biobank did not contain a high prevalence of dilated or dysfunctional RV measurements, we fine-tuned the model on a clinical cohort of diverse patients in a large urban health system with five hospitals (MSH_original_; The Mount Sinai Health System, New York). We queried our cMRI reporting software (Precession, Intelerad, Montreal, Canada) for studies performed from April 04, 2012 to November 22, 2022 in patients ≥ 18 years old, and collected clinically-indicated cMRI reports paired with ECGs within two-weeks before or two-weeks after cMRI exam. The MUSE Cardiology Information System (GE, Boston, MA) ECG database was queried for relevant patient medical record numbers, filtered by date of acquisition, and exported as .xml files. We excluded patients with congenital heart disease as an indication for MRI, or without a recorded ECG within the inclusion timeframe. Like the UKBB dataset, we excluded entries with RVEF values < 10% or > 80%. We extracted clinically measured RVEDV and RVEF from reports. MRI is typically performed on a 1.5T scanner using balanced-SSFP sequences with manual contouring of the cine short axis stack to include RV papillary muscles within the blood pool for the RV, though all reports were included. Left ventricular volume (contoured to exclude the papillary muscles from the blood pool) and ejection fraction were also collected. Patient age at cMRI, sex, indication for cMRI, cardiac rhythm at cMRI, weight, and height were also collected from the report.

### ECG preprocessing

Our ECG preprocessing pipeline is described in detail previously^14^. ECGs in both the UKBB and MSH datasets were extracted as XML files. We included eight channels: leads I, II, and V1-V6 as the other leads are linear transformations without extraneous information. We excluded low-quality ECG recordings quantified by amplitude greater than +/-3 SD of the population mean. ECGs were filtered to remove noise, restricted to the first five seconds to ensure all patients had equivalent data, and plotted to images.

### Model construction

The model input was the cMRI-paired 12-lead ECG. The model outputs of interest were numerical RVEF (%) and RVEDV/body surface area (mL/m^2^) (i.e., supervised learning for regression tasks) as well as these values dichotomized into pathological thresholds of RV EF < 40% and RVEDV ≥ 120 mL/m^2^ (i.e., supervised learning for classification tasks). In the UKBB study protocol only one ECG was obtained per subject and cMRI, which was randomly split into 80% training and 20% testing subsets. For the MSH_original_ dataset, as multiple ECGs per cMRI could occur within the inclusion time frame, patients were group shuffle split into 80% train and 20% test groups. This method ensured that no patient was represented both in the test and train set, but allowed multiple ECGs obtained in the inclusion period for a patient to be considered in the train set. For the MSH_original_ test set, only one subject with one paired ECG closest to the time of cMRI was included.

We selected a DenseNet-201 two-dimensional convolutional neural network architecture which had previously been trained on > 700,000 ECGs^14^ as initialization parameters. We trained the network using the Adam optimizer with cross-entropy as the loss function for classification and mean absolute error (MAE) for regression tasks, respectively. We first trained on the UKBB dataset as a way of training features specific to the right ventricle, but the dataset has a low prevalence of RV pathology. We then fine-tuned the model to predict outcomes in the MSH cohort which had a higher outcome prevalence to learn features specific to RV pathology. Fine-tuning a neural network refers to the process of using pre-trained parameters (as opposed to random parameters) from a related task as a starting point for training on a new task or dataset. This may potentially improve model performance on smaller datasets since the network may have already learned the basic features needed for the new task.

### Model evaluation

We evaluated model performance with area under the receiver operating curve (AUROC) and area under the precision recall curve (AUPRC) for classification tasks. Precision-recall curves are generated by plotting the precision (i.e., the positive predictive value) against the recall (i.e., the sensitivity) over the range of predictions generated. AUPRC, by emphasizing the ability of the classifier to predict outcomes, is useful to evaluate the performance of a classifier in the setting of class imbalance where outcomes are more important to detect than non-outcomes^15^. Unlike AUROC, AUPRC depends on the baseline prevalence of disease and therefore can be interpreted as the ability of the classifier to identify a group “enriched” for disease over the baseline. We also evaluated classification model performance in key subgroups including sex, LV dysfunction (defined as LVEF < 50%), arrhythmia, obesity (BMI ≥ 30), and age ≥ 60 years. We performed AUROC comparisons and tested for effect modification between subgroups using the method of DeLong^16^. For regression tasks, we generated observed-versus-expected scatterplots and evaluated agreement with R^2^ calculations, Lin’s concordance correlation coefficient, and Bland-Altman plot analysis. We calculated mean absolute error (MAE) of the models with 95% confidence interval through 500 bootstrap iterations. We also performed saliency mapping using the GradCAM library to highlight the regions of a given ECG input image that were most associated with a prediction.

### Temporal validation

We performed temporal validation (MSH_validation_) of the final classification models in the period of 11/23/2022 to 03/21/2023 from a cohort of patients with clinically indicated cMRI and paired ECG within +/-14 days of cMRI like the MSH_original_ dataset. Patients with prior ECG or cMRI included in the MSH_original_ dataset were excluded. AUROC and AUPRC metrics were reported as above.

### Survival Analysis for Composite Outcome

We reviewed medical records of patients included within the MSH_original_ test set to ascertain date of last known follow-up, mortality, and date of heart transplant if performed. We performed survival analysis with Cox proportional hazards models to evaluate the association between predicted numerical RVEF and the combined outcome of death or heart transplantation. We included other co-variates including MRI-quantified LVEF, age, sex, and self-reported race/ethnicity. We tested proportional hazards assumptions by evaluating the relationship between scaled Schoenfeld residuals and time. A p value < 0.05 was considered statistically significant.

### Software

Model training was performed in Python (3.8) with pandas, numpy, scipy, scikit-learn, PyTorch, torchvision, PIL, matplotlib, seaborn, and GradCAM packages. Data visualization and descriptive statistics were performed in R and Stata. Continuous metrics are reported as mean (standard deviation). Code for neural network training will be made publicly available at https://github.com/akhilvaid/RVSizeFunction.

## Results

### Demographic and clinical information

We included 42,938 patient MRI-ECG pairs from UKBB after quality control. Mean age was 64.7 years, 48% were male, 96.5% were white, and the most common comorbidities were hypertension (22.5%) and hyperlipidemia (17.3%). Heart surgery and history of myocardial infarction were reported in 1.8% and 1.4%, respectively. The mean RVEF was 57% (SD 6.3), with RV dysfunction (RVEF ≤ 40%) present in 425 (1.0%). Mean RVEDV was 82 (15.6) mL/m^2^ with RV dilation (RVEDV ≥ 120 mL/m^2^) in 680 (1.6%).

The MSH_original_ dataset consisted of 3,019 patients that were split 80-20% into train and test cohorts. The training set consisted of 2,415 patients and 13,673 ECG recordings meeting the inclusion criteria. The test set contained 604 patients each with one paired cMRI and ECG. Test set mean age was 56 (17) years and cardiomyopathy was the predominant indication for cMRI (80%). Prevalence of RV dysfunction was 18%, and prevalence of RV dilation was 11%. Demographic, clinical, and MRI characteristics of the MSH_original_ cohorts split by test and train groups are listed in Table 1.

**Table 1:** Demographic and clinical characteristics in MSH dataset

|  | MSH<sub>original</sub><br>Train set<br>(n=2,415) | MSH<sub>original</sub><br>Test set<br>(n=604) | MSH<sub>validation</sub><br>Temporal<br>Validation<br>(n=115) |
| --- | --- | --- | --- |
| Age, years (mean (sd)) | 56.0 (16.4) | 55.9 (17.0) | 56.3 (16.1) |
| Sex, Female (%) | 865 (37.5%) * | 209 (36.3%) ** | 45 (39.8%) *** |
| Sex, Male (%) | 1,550 (62.5%) | 395 (63.7%) | 70 (60.2%) |
| Race/Ethnicity: White | 995 (41.2%) | 258 (42.7%) | 41 (35.7%) |
| Black | 476 (19.7%) | 101 (16.7%) | 28 (24.4%) |
| Other/Unknown | 944 (39.1%) | 245 (40.6%) | 46 (40%) |
| Body Surface Area, m<sup>2</sup><br>(mean (sd)) | 1.95 (0.29) | 1.96 (0.27) | 1.95 (0.29) |
| BMI | 27.7 (6.4) * | 27.6 (5.8) ** | 27.6 (6.2) |
| BMI ≥ 30 kg/ht<sup>2</sup> | 663 (28.5%) * | 152 (26%) ** | 36 (31.3%) |
| Indication for cMRI: Cardiomyopathy | 1,959 (81.1%) | 485 (80.3%) | 86 (74.8%) |
| Abnormal ECG | 183 (7.6%) | 56 (9.3%) | 1 (0.9%) |
| Tumor | 107 (4.4%) | 22 (3.6%) | 8 (7.0%) |
| Valvular function | 20 (0.8%) | 5 (0.8%) | 0 (0%) |
| Other/unknown | 146 (6.0%) | 36 (6.0%) | 20 (17.4%) |
| Rhythm at cMRI: Normal Sinus | 1,957 (91.1%) * | 476 (90.5%)** | 106 (95.5%)*** |
| Sinus with ectopic beats | 97 (4.5%) | 25 (4.8%) | 2 (1.8%) |
| Atrial fibrillation/flutter | 78 (3.6%) | 21 (4.0%) | 1 (0.9%) |
| Other | 15 (0.7%) | 4 (0.7%) | 2 (1.8%) |
| Admitted as inpatient at time of MRI | 863 (35.7%) | 217 (35.9%) | 45 (39.1%) |
| LVEF (median [IQR]) | 53 [36-61]* | 53 [36-62] | 52[34-63] |
| LVEF <50% | 1016 (42%)* | 261 (43.4%) | 44 (38.3%) |
| RVEF (median [IQR]) | 54 [46-61] | 53 [46-60] | 54 [47-59] |
| RVEF ≤40% | 411 (17.0%) | 109 (18.0%) | 18 (15.7%) |
| LVEDVi (median [IQR]) | 84 [69-110]* | 84 [70-108]** | 85[65-107] |
| RVEDVi (median [IQR]) | 78 [63-95]* | 80 [64-99]** | 70 [58-81] |
| RVEDVi ≥120 | 202 (8.7%) | 62 (10.6%) | 5 (4.3%) |

\*Train set missing data: Rhythm at cMRI n=268; Patient sex n=108; LVEDVi n=91; BSA, BMI, RVEDVi n=87; LVEF n=4
\*\*Test set missing data: Rhythm at cMRI n=78; BSA, BMI, LVEDVi, RVEDVi n=20 missing, LVEF n=3 missing, Patient Sex n=28 missing.
\*\*\*Temporal validation set missing data: Patient rhythm n=4; patient sex n=2

The MSH_validation_ group consisted of 115 patients, each with one paired ECG and cMRI. Demographic and clinical characteristics were concordant with MSH_original_ group, except for smaller RVEDV measurements and a lower prevalence of RV dilation (n = 5, 4.3%). However, presence of RV systolic dysfunction was similar (n = 18, 15.7%). Clinical and demographic information of the temporal validation cohort is shown in Table 1.

### RVEF classification models

Model performance characteristics to classify RV systolic dysfunction (defined as RVEF ≤ 40%) in UKBB and MSH datasets are shown in Figure 1. In UKBB (baseline prevalence 1.0%), AUROC was0.86 [0.80-0.90] and AUPRC was 0.17 [0.11-0.26]. In the MSH_original_ dataset (baseline prevalence 18.0%), AUROC was 0.81 [0.77-0.86] and AUPRC was 0.59 [0.49-0.69]. Performance between clinically relevant subgroups in the MSH_original_ test dataset is shown in Table 2. RVEF model AUROC did not differ between any tested subgroup, including those with and without LV dysfunction.

**Table 2:** Classification Models, MSH subgroup analysis

|  |  |  | Model: | p* | Model | p* |
| --- | --- | --- | --- | --- | --- | --- |
|  |  | n | MSH RVEDV<br>classification |  | MSH RVEF<br>classification |  |
| Overall |  | 604 | 0.81 [0.76-0.86] |  | 0.81 [0.77-0.86] |  |
| Subgroup | Female | 209 | 0.72 [0.62-0.81] | 0.40 | 0.79 [0.69-0.90] | 0.46 |
|  | Male | 395 | 0.77 [0.70-0.83] |  | 0.84 [0.79-0.89] |  |
| | Age ≥ 60 years | 272 | 0.67 [0.57-0.76] | 0.015 | 0.80 [0.72-0.87] | 0.58 |
|  | Age < 60 years | 332 | 0.81 [0.74-0.87] |  | 0.82 [0.76-0.89] |  |
|  | Arrhythmia | 50 | 0.74 [0.53-0.94] | 0.56 | 0.81 [0.67-0.94] | 0.69 |
|  | Non-arrhythmia | 476 | 0.80 [0.74-0.86] |  | 0.84 [0.79-0.89] |  |
|  | LV EF < 50% | 261 | 0.87 [0.81-0.94] | 0.024 | 0.74 [0.68-0.81] | 0.93 |
| | LV EF ≥ 50% | 340 | 0.75 [0.67-0.83] | | 0.75 [0.53-0.98] | |
| | BMI ≥ 30 kg/ht<sup>2</sup> | 152 | 0.80 [0.75-0.88] | 0.79 | 0.88 [0.73-0.86] | 0.053 |
|  | BMI < 30 kg/ht<sup>2</sup> | 432 | 0.80 [0.68-0.91] |  | 0.79 [0.73-0.86] |  |
|  | Race: White | 258 | 0.83 [0.74-0.92] | 0.83 | 0.82 [0.74-0.91] | 0.90 |
|  | Race: Black | 101 | 0.78 [0.64-0.92] |  | 0.82 [0.74-0.91] |  |
|  | Race: Other/Unknown | 245 | 0.82 [0.75-0.89] |  | 0.79 [0.72-0.88] |  |

\*p-value for comparison of AUROC curves between subgroups

**Figure 1.**
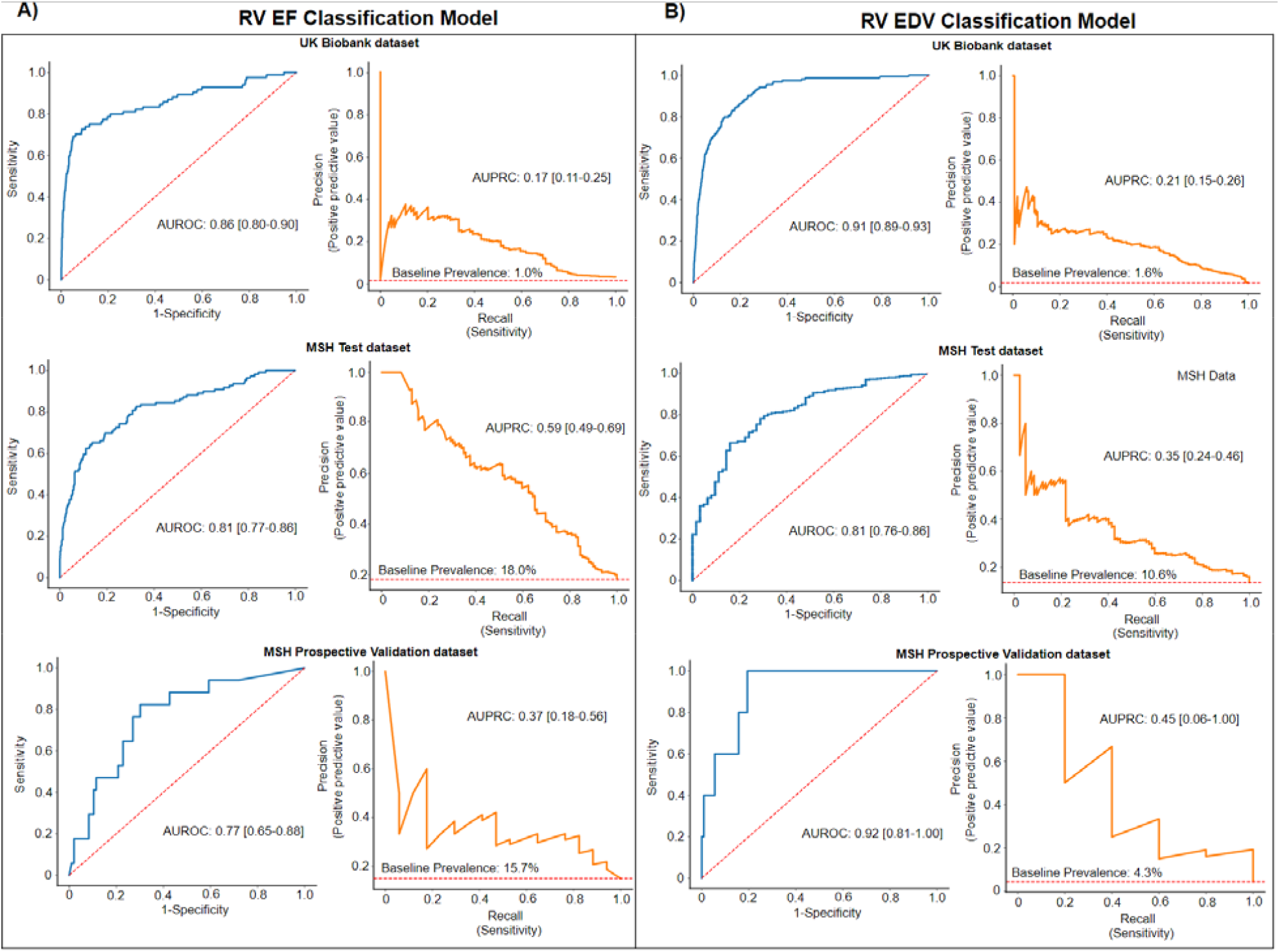
Classification results for Abnormal Right Ventricular Ejection Fraction and End-Diastolic Volume Caption: Receiver operating characteristic curves (Blue) and Precision-Recall curves (Orange) to classify RV systolic dysfunction (A,left) and RV dilation (B, right) in UK Biobank dataset (top), MSH dataset (middle), and MSH prospective validation dataset(bottom). Red-dashed lines on ROC curves represents no model skill, and on PR curves represents baseline incidence of disease. AUROC: Area under the receiver operating curve, AUPRC: Area under the precision recall curve.

### RVEDV classification models

Model performance to classify RV dilation (defined as RVEDV ≥ 120 mL/m^2^) are shown in Figure 1b. In UKBB (baseline prevalence 1.6%), AUROC was 0.91 [0.89-0.93] and AUPRC was 0.21 [0.15-0.26]. In the MSH_original_ dataset (baseline prevalence 10.6%), AUROC was 0.81 [0.76-0.86] and AUPRC was 0.35 [0.24-0.46]. Classification performance between clinically relevant subgroups in the MSH dataset are shown in Table 2. RVEDV model performance differed in patients ≥ 60 vs < 60 years (AUROC 0.67 vs. 0.81; p = 0.015), and those with LV systolic dysfunction vs normal LV function (AUROC 0.87 vs. 0.75; p = 0.024).

### Prospective Validation

The final RVEDV and RVEF classification models were further tested in the MSH_validation_ group, a temporal validation dataset consisting of ∼4 months of data collected after development of the final models. For RV systolic dysfunction classification, model AUROC was 0.77 [0.65-0-88] and AUPRC 0.37 [0.18-0.56]. For RV dilation classification, model AUROC was 0.92 [0.81-1.00] and AUPRC 0.45 [0.06-1.00].

### RVEDV and RVEF Regression models

We developed models to predict the numerical value of RVEDV and RVEF. They were subsequently fine-tuned in the MSH dataset with test set regression results shown in Figure 2. For RVEF prediction the mean absolute error between predicted and expect RVEF was clinically reasonably low at 7.8%, the R^2^ = 0.360 for the linear fit between the two measures, and there was moderate agreement (concordance correlation coefficient= 0.57). Bland-Altman analysis confirmed that the average mean difference between measures was low, but limits of agreement were wide (mean difference -0.4%, 95% LOA [-20.4, 19.7]). The errors were non-normally distributed with ECG overestimating cMRI RVEF at low (pathological) values and ECG underestimating cMRI RVEF at higher values. In the RVEDV model, the mean absolute error was 17.6 mL/m^2^, R^2^=0.250, concordance correlation coefficient was 0.43. Like the RVEF models, Bland-Altman analysis confirmed low average mean difference but wide limits of agreement (mean difference -2.2 mL/m2, 95% LOA [-49.9, +45.4]) and non-normally distributed error. ECG overestimated cMRI RVEDV at low values and ECG underestimating cMRI RVEDV at higher (pathological) values.

**Figure 2.**
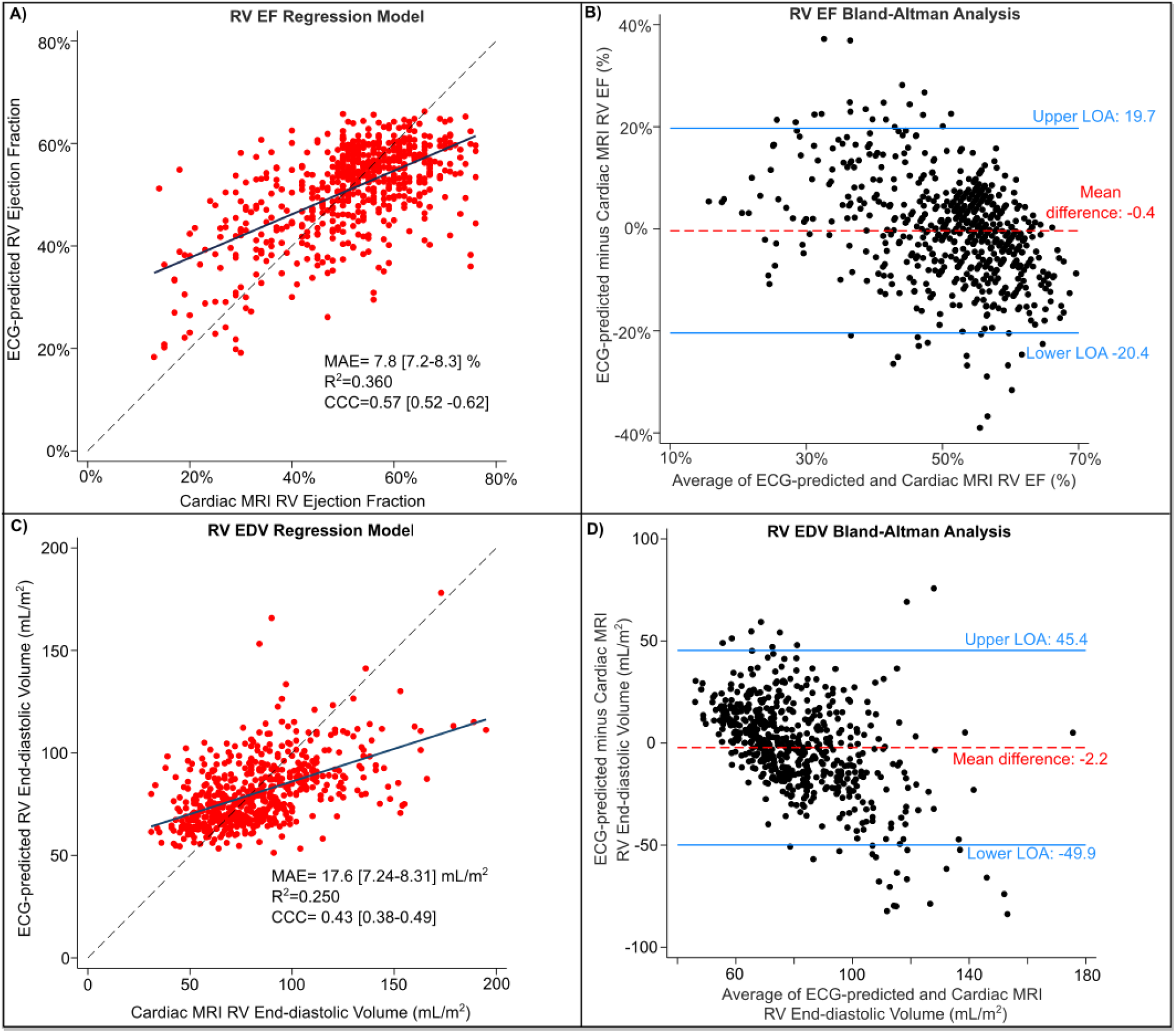
Regression Models, MSH dataset Caption: MSH_original_ regression model with ECG-predicted versus cMRI-expected scatter plots to predict numerical RVEF (Box A) and numerical RVEDV (Box C). Scatter plots plotted with line of perfect concordance (dashed black line) and linear best-fit line (solid blue). Bland-Altman analysis of RVEF (Box B) and RVEDV (Box D) with mean difference (red dashed line) and upper/lower limits of agreement (blue solid line).

### Saliency Mapping

RV dilation and RV dysfunciton classification model saliency mapping examples from MSH_original_ test set are shown in Figure 3. Qualitative review of several saliency mapping examples suggested that P waves, and QRS complexes in leads II, V1, and V5/6 are particularly influential.

**Figure 3.**
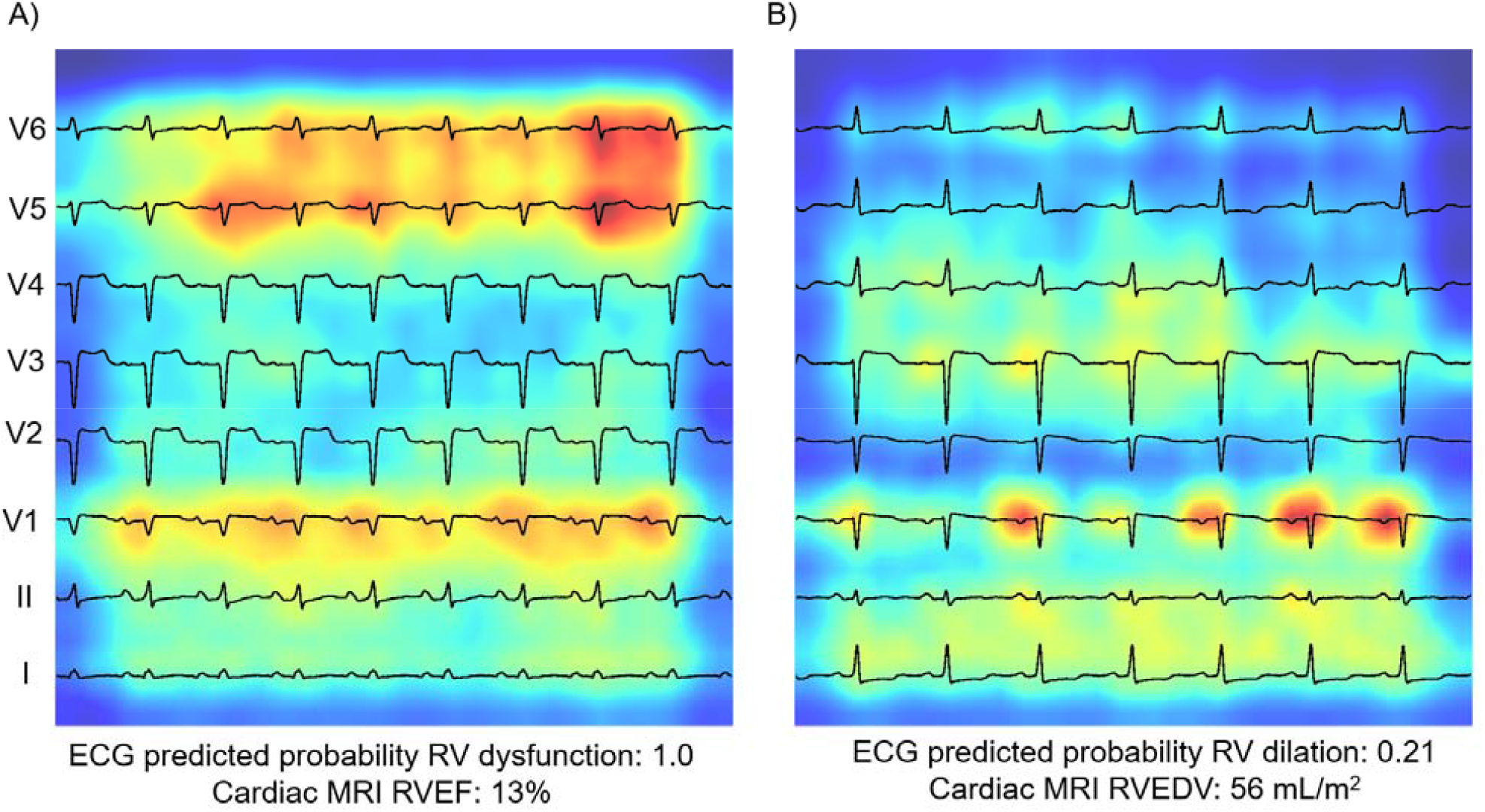
Saliency Mapping to Identify Key Areas of the ECG relevant to the prediction. Caption. Saliency mapping ECG examples from the MSH_original_ test set. Leads are arranged from bottom to top: lead I, II, V1-V6. Increasing shades of red indicate increasingly influential pixel in the model. Figure A) shows a true positive example of RV dysfunction. The P and QRS portions of V1, V5, and V6 are particularly influential. Figure B) shows a true negative example of RV dilation. QRS complex of lead I and P-wave in V1 are particularly influential

### Survival analysis

Follow-up information was available for 575/604 (95%) patients in the MSH_original_ test cohort with a median follow-up time was 2.27 [IQR 0.67-5.03] years. The combined outcome of death or heart transplantation occurred in 65/604 (10.8%, n = 59 deaths and n = 6 heart transplants) patients. Survival analyses are shown in Table 3 for the combined outcome of death and heart transplantation. Lower ECG-predicted RVEF was associated with outcome risk. In a model including MRI-quantified LVEF and age, ECG-predicted RVEF was independently associated with increased risk of death/transplant (Hazard Ratio 1.37 for each 10% decrease in RVEF, p = 0.046). Other demographic variables including sex, race/ethnicity and BMI were not significant in bivariable analysis, and no significant interaction was identified between LVEF and RVEF.

**Table 3:** Survival analysis for freedom from death/ heart transplant, bi- and multivariable analyses

| Cox Proportional Hazards Models | Model Covariates | Hazard Ratio [95%CI] | P |
| --- | --- | --- | --- |
| Model 1: | Predicted RVEF (every 10% decrease) | 1.41 [1.12-1.75] | 0.003 |
| Model 2: | MRI-quantified LVEF (every 10% decrease) | 1.18 [1.02-1.33] | 0.026 |
| Model 3*: | Predicted RVEF (every 10% decrease) | 1.37 [1.01-1.85] | 0.046 |
|  | MRI-quantified LVEF (every 10% decrease) | 1.05 [0.88- 1.25] | 0.55 |
|  | Age (years) | 1.04 [1.02-1.05] | <0.001 |

\*interaction between ECG predicted RVEF and MRI-LVEF was not significant. Tested Race/Ethnicity, sex, BMI, all of which were not significant in bivariable analysis.

## DISCUSSION

In summary, we utilized deep learning on 12-lead ECGs to quantify RVEDV and RVEF based on paired cMRI as reference standard. We first developed this model in a large cohort of mostly healthy participants in the UKBB registry, then further fine-tuned the model in a smaller cohort of clinically indicated paired ECG and cMRI samples from MSH. Finally, we prospectively validated the model in the subsequent ∼4 months after model development. The classification models to detect RV dilation (RVEDV ≥ 120 ml/m2) and RV dysfunction (RVEF ≤ 40%) have good performance in a clinical cohort. Regression models demonstrated moderate predictive ability of ECG to numerically estimate cMRI-measured RVEDV and RVEF, and ECG-predicted RVEF was associated with worse short-term freedom from death/heart transplantation. Saliency mapping suggested P-wave and QRS complexes of leads II, V1, and V5/V6 as particularly important. This represents a significant advancement in prediction of RV enlargement by 12-lead ECG which is known to be insensitive by traditional methods (e.g. voltage criteria)^17^, and this work is the first to show that RV volumetric markers of dilation and dysfunction can be predicted from ECG.

We find the classification models to be more clinically applicable, as they provide good test performance characteristics at clinically relevant thresholds. As an example of the clinical applicability of the current models: at a baseline prevalence of RV dysfunction of ∼18% in the MSH dataset, examination of the AUPRC curve reveals that at a prediction threshold recall (sensitivity) of ∼50%, the precision (positive predictive value) is ∼60%. In other words, a simple 12-lead ECG in this population could alert the practitioner to half of all cases of RV dysfunction in the population, and of cases identified, over half would have true RV dysfunction. In contrast to the classification model performance, our regression model performance is less clinically applicable at this time as Bland-Altman analysis suggested there is high bias at the extremes of measurement (which are the pathological cases) and overall wide limits of agreement between ECG and MRI. However, the fact that a correlation is detected at all is itself a novel breakthrough in DL-ECG analysis. Additionally, ECG-predicted RVEF was associated with combined outcome of death/heart transplant independent of demographics and LV function in short term follow-up, which demonstrates the potential clinical utility of this algorithm for prediction of patient outcome.

DL-ECG models have been shown to predict left ventricular systolic dysfunction, elevated left ventricular mass, and primarily left-sided structural heart disease^14,18-20^. The right-heart is largely uninvestigated in this field. We have previously shown that DL-ECG analysis can detect qualitative RV systolic dysfunction or RV dilation by two-dimensional echocardiography^14^, however the limitations of two-dimensional echocardiography in quantitative assessment of the right ventricle are well described.

Our model represents an advancement over this prior work through training on a diverse group of datasets with paired reference standard cMRI RV measurements, thus allowing us to quantify RV size and systolic function by an important clinical metric. Although further work is needed, DL models may eventually be incorporated into on-cart analysis to provide a useful screening tool for RV health that is validated against reference standard metrics.

In the MSH cohort, LV and RV dysfunction were often found together, but our model performed well in the subgroups with and without concomitant LV dysfunction. This suggests that our model can distinguish RV systolic dysfunction independently of LV dysfunction. Similarly, equivalent performance in those with and without arrhythmia suggests the model recognizes RV systolic dysfunction independently of cardiomyopathy related to abnormal rhythm. Our subgroup analysis suggests the classification models have robust test characteristics across subgroups with described differences in ECG morphology including age, BMI, sex, and race.

Although the depolarization and repolarization patterns on the 12-lead ECG are dominated by the higher myocardial mass of the left ventricle, prior work has described crude correlations between ECG pattern and right ventricular systolic function^21-23^, which provide a potential scientific rationale for our findings. In our study, saliency mapping demonstrated the importance of lead V1 in evaluating the right ventricle, which may be physiologically explained by its proximity to the right atrium and right ventricle. Further study to differentiate how and why these models can RV dysfunction independently from LV dysfunction are needed to enhance the explainability of DL-ECG RV models.

This work should be viewed in the light of some limitations. First, model training and fine-tuning was performed in only two datasets in two settings, which may limit the generalizability of these models to other relevant cardiac conditions. The overwhelming majority of patients in the MSH cohort had indication of cardiomyopathy for cMRI, and a higher-than-expected number of patients were admitted to the hospital at time of MRI. This was likely in part due to the tight inclusion period imposed on paired MRI-ECG samples to ensure ECGs were valid representations of RV health. However, as MRI is more commonly a scheduled outpatient procedure, it is possible that other important patient subgroups were excluded. Therefore, external validation in other populations, for instance in pulmonary hypertension and congenital heart disease where ECG patterns are likely to differ, will ensure the generalizability and clinical applicability of these models. Second, we trained models on the large UKBB dataset, in which RV volumes were automatically contoured through a cMRI segmentation algorithm. This method may be more prone to error and systematically biased compared to clinician labeling, and this method differed from the MSH cohort in which we derived RV measurements from clinician-verified cMRI reports. We finetuned the model on MSH data to account for the differences in distribution between datasets, but it is possible that we might have obtained better performance if the UKBB dataset were labeled by clinicians. However, this is infeasible due to the large number of studies (> 40,000) in UKBB. Third, we used ECG information plotted to images and utilized a 2-dimensional CNN architecture in modeling. Our rationale for this is that not all centers may store ECG data as 1D vectors, making an image-based approach more universally acceptable. Additionally, a 2D approach allows us to take advantage of availability of 2D CNN models pretrained on millions of images. It is possible that plotting to images may result in loss of information and complicate model training, however others have suggested superior performance of image-based over signal-based ECG input methods^24^. Further research to compare and define the benefits of different input formats and model architectures is required. Finally, our regression models demonstrate the potential of deep learning analysis to derive unseen functional data from the 12-lead ECG, however currently they exhibit clinically unacceptable limits of agreement with cMRI. Steps to improve prediction may require experimentation with different model architectures, training on additional pathological cases, and resampling techniques. As these models improve, research into the usefulness of serial DL-ECG analysis to trend RV size or function should be explored, as trending of RV metrics over time is an important clinical application of this technology that may reduce the need for serial cMRI.

In summary, we developed and validated a deep learning algorithm to predict RV dilation or systolic dysfunction as defined by reference standard cMRI metrics. This unique method can provide novel RV quantification from a simple and globally available tool. These prediction tools may be useful in decreasing the need for costly cMRI, with implications for disease screening, risk stratification, and progression of disease. Future work to improve accuracy of prediction, to validate findings in heterogeneous clinical populations, and clinical trials to show improved workflow and outcomes are required.

## Data Availability

Code for neural network training will be made publicly available at https://github.com/akhilvaid/RVSizeFunction.

## Abbreviations

RV: Right Ventricle
RVEDV: Right ventricular end-diastolic volume
RVEF: Right ventricular ejection fraction
ECG: Electrocardiogram
DL: Deep learning
cMRI: cardiac MRI
MAE: Mean absolute error

